# Adherence to protective measures among health care workers in the UK; a cross-sectional study

**DOI:** 10.1101/2020.07.24.20161422

**Authors:** Louise E. Smith, Danai Serfioti, Dale Weston, Neil Greenberg, G James Rubin

**Affiliations:** King’s College London, Institute of Psychiatry, Psychology and Neuroscience; NIHR Health Protection Research Unit in Emergency Preparedness and Response; Public Health England, Behavioural Science Team, Emergency Response Department Science and Technology

**Keywords:** COVID-19, personal protective equipment, hand hygiene, physical distancing, government measures

## Abstract

Healthcare workers (HCWs) are frontline responders to emergency infectious disease outbreaks such as COVID-19. We investigated factors associated with adherence to personal protective behaviours in UK HCWs during the COVID-19 pandemic using an online cross-sectional survey of 1035 healthcare professionals in the UK. Data were collected between 12^th^ and 16^th^ June 2020. Adjusted logistic regressions were used to separately investigate factors associated with adherence to use of personal protective equipment, maintaining good hand hygiene, and physical distancing from colleagues. Adherence to personal protective measures was suboptimal (PPE use: 80.0%, 95% CI [77.3 to 82.8], hand hygiene: 67.8%, 95% CI [64.6 to 71.0], coming into close contact with colleagues: 74.7%, 95% CI [71.7 to 77.7]). Adherence to PPE use was associated with having adequate PPE resources, receiving training during the pandemic, lower perceived fatalism from COVID-19, higher perceived social norms and higher perceived effectiveness of PPE. Adherence to physical distancing was associated with one’s workplace being designed, using markings to facilitate physical distancing and receiving training during the pandemic. There were few associations with adherence to hand hygiene. Findings indicate HCWs should receive training on personal protective behaviours to decrease fatalism over contracting COVID-19 and increase perceived effectiveness of protective measures.

## INTRODUCTION

Healthcare workers (HCWs) are frontline responders to infectious disease outbreaks such as COVID-19. Without adequate protective measures, there is potential for the rapid spread of disease within hospital, or other care settings, among both patients and staff and onwards to the general population. To help prevent this, a series of protective measures have been introduced, including the use of personal protective equipment (PPE: surgical masks, gloves, aprons or gowns and eye or face protection), maintaining good hand hygiene, and physical distancing from others.^1-3^ Public Health England guidance indicates that staff should be trained in the correct use of PPE during COVID-19.^1^ In spite of these measures, evidence suggests that the incidence of COVID-19 infection is elevated in patient-facing UK HCWs and resident-facing social care workers than those not direct contact roles.^4, 5^ This is likely to be due to increased exposure to the virus, and highlights the importance of adherence to protective measures among staff.

Studies conducted before the COVID-19 pandemic have highlighted factors that help HCWs adhere to personal protective behaviours such as PPE use, hand hygiene and physical distancing. A recent rapid review of adherence to infection control measures during emerging infectious disease outbreaks (56 papers, two relating to COVID-19) found evidence that better adherence was associated with higher perceived effectiveness of infection control measures, higher perceived risk of the illness.^6^ Poor adherence was associated with concerns about shortages of PPE, inadequate guidance for use of PPE, perceived negative impact of PPE on a HCWs ability to communicate with patients, and seeing other colleagues not adhering to PPE.

Hand hygiene in hospitals is also far from perfect, with one review estimating median compliance rates of 40%.^7^ However, there are no data investigating adherence to hand hygiene in UK hospitals during COVID-19. Previous research indicates that poorer hand hygiene was found in intensive care units compared to other settings, in physicians than nurses, and in those with a greater workload.^7^ Better hand hygiene was associated with seeing colleagues and clinical leaders adhering to hand washing guidance, increased motivation to protect oneself and one’s family from illness, adequate facilities, and having an organisational culture that encourages good hand hygiene; hand hygiene was not adhered to when it delayed emergency or lifesaving care for a patient.^8^

We used a cross-sectional survey of healthcare professionals during the COVID-19 pandemic to investigate factors associated with adherence to use of PPE, good hand hygiene, and physical distancing in the workplace. We also investigated factors associated with total number of outings in the last 24 hours (supplementary materials).

## METHOD

### Design

We commissioned the market research company YouGov to carry out this cross-sectional survey, between 12^th^ and 16^th^ June 2020.

### Participants

Participants (n=1035) were recruited from YouGov’s online research panel (n=1,000,000+ UK adults) and were eligible for the study if they were aged eighteen years or over, lived in the UK and worked in the healthcare sector. Quota sampling was used, based on occupational group. Of 1601 people who began the survey, 1047 completed it. Twelve participants were not included in the sample following quality control procedures including review of open-ended responses, suspiciously speedy completion of the survey (“speeding”) or providing identical answers to multiple consecutive questions (“straight-lining”). Participants were reimbursed in points (equivalent to approximately 50p) which could be redeemed as cash, gift vouchers or charitable donations.

### Study materials

Full survey materials are available in the supplementary materials.

#### Outcome measures

Questions about personal protective behaviours at work were only asked to those who had been to work in the last six weeks.

We asked participants about their use of PPE (mask, gloves, apron or gown, and face or eye protection) the most recent time they were at work. Separate questions were asked for each component of PPE. Possible answers included: managing to wear PPE every time you were meant to; not managing to wear PPE every time you were meant to and wearing PPE even if you were not supposed to.

Participants were asked if they had managed to wash their hands with soap and water for 20 seconds or apply hand gel: every time they needed to at work; as soon as they got to work; before eating at work; as soon as they got home; and whether they had a shower before leaving work or as soon as they got home.

We asked participants if they had been in close contact with someone else who works in the health sector (with close contact defined as coming within 2 metres for 15 minutes or more): during a team meeting; on a ward/unit/clinical area when not wearing PPE; in a break room, café or canteen; or in a corridor. We also asked if they had direct physical contact with someone else who works in the health sector.

#### Workplace environment

Questions about the workplace environment were only asked to those who had been to work in the last six weeks.

Participants were asked about the facilities at their workplace, including whether: facilities were available to make it easy to wash their hands when they got into work; their workplace had clear markings to help them stay 2 metres away from others; their workplace was designed to make it easy for them stay 2 metres away from others; they had received adequate health and safety training in their workplace during the COVID-19 pandemic (e.g. on correct use of PPE and physical distancing); they were given all the correct PPE they needed to do their job safely; and they had enough information about which item of PPE to use and when to use it. Responses were given on a five-point Likert scale from “strongly agree” to “strongly disagree”. We also asked participants if they was an instance in their last week at work that they could not wash their hands because a sink was broken; or because there was no soap or paper towels, or an empty gel dispenser (yes / no).

Participants were asked how easy or difficult they found it to keep 2 metres away from other people in their workplace while: eating in canteens; during rest breaks; when saying hello or goodbye to colleagues; when moving between areas (e.g. in corridors); and when carrying out work that does and does not involve patient contact. Answers were given on a five-point Likert-type scale from “very easy” to “very difficult”.

#### Psychological and situational factors

We asked participants to rate to what extent they thought that COVID-19 posed a threat to people in the UK and themselves personally on a five-point Likert type scale from “no risk at all” to “major risk”.

We asked participants whether they thought they “had, or currently have, coronavirus”. Possible answers were “I have definitely had it or definitely have it now”, “I have probably had it or probably have it now”, “I have probably not had it and probably don’t have it now”, and “I have definitely not had it and definitely don’t have it now”.

Participants were asked if they had experienced symptoms “in the past seven days” and if someone else in their household had experienced symptoms “in the past fourteen days” from a list of thirteen symptoms including cough, high temperature / fever and loss or change to their sense of smell or taste.^9, 10^

We measured perceived credibility of information from the NHS about PPE using an adapted form of the Meyer Credibility Scale.^11^

We asked participants to rate twenty-one perception statements on a five-point Likert scale from “strongly disagree” to “strongly agree”. Statements included perceived effectiveness of different items of PPE and physical distancing; perceived social pressure from colleagues to maintain physical distancing and wear PPE; and perceived safety from COVID-19 at work, home and when out and about (see supplementary materials for full item list). Perception questions were only asked to those who had been to work in the last six weeks.

We also asked participants how they would prefer to get updates related to PPE.

#### Personal and occupational characteristics

We asked participants to report their age, sex and region of their place of work.

We asked participants for their occupational group, work setting. We also asked participants if they had face-to-face contact with patients or service users as part of their job, and how frequently they came into contact with patients with COVID-19 or staff who worked closely with patients with COVID-19.

### Ethics

Ethical approval for this study was granted by the King’s College London Research Ethics Committee (reference: LRS-19/20-19184).

### Patient and public involvement

To inform survey materials, we carried out brief telephone interviews with a small group of clinical and administrative staff working in healthcare settings (n=5).

### Power

A sample size of 1035 allows a 95% confidence interval of plus or minus 3% for the prevalence estimate for each survey item.

### Analysis

#### Recoding of variables

We created a single binary variable indicating whether participants fully adhered to wearing PPE (mask, gloves, apron or gown and eye or face protection) or not. For these items, we coded people who wore items of PPE even when they did not need to as adherent.

We created a binary variable indicating whether participants had been in close contact with a colleague while at work.

We recoded whether people thought they had had COVID-19 or thought they had it now into, and presence of household symptoms into separate binary variables. Presence of symptoms was defined as a participant reporting that they had experienced cough, a high temperature / fever, or loss or change to their sense of smell or taste in the last seven days, or if a member of their household had experienced cough, a high temperature / fever, or loss or change to their sense of smell or taste in the last fourteen days.^9^ Under UK guidance at the time, either of these events should have resulted in the participant being required to not leave their home at all for a minimum of seven days.

We recoded perceived ease of maintaining physical distancing in different situations in the workplace and effectiveness of wearing PPE (face mask and gloves around patients, and face mask around colleagues) into separate continuous variables.

For all variables, unless stated otherwise, we coded answers of “don’t know” as missing data.

#### Analyses

We used a series of logistic regressions to investigate univariable associations between (a) total adherence to the use of PPE, (b) hand washing when needed at work, and (c) close contact with colleagues at work. We investigated associations with personal and occupational characteristics, work environment, and psychological and situational factors. We ran a second set of logistic regressions controlling for personal and occupational characteristics (sex, age, region of place of work, sector, work setting, and face-to-face contact with patients or service users).

Data were weighted by occupation group in the NHS workforce.

#### Sensitivity analyses

Due to the large number of analyses run on each outcome (PPE, n=30; hand hygiene, n=23; physical distancing, n=27), we applied a Bonferroni correction to our results *p*≤.002. Those meeting this criterion are marked by a double asterisk (**) in the tables.

## RESULTS

Only results of adjusted analyses are reported narratively; unadjusted results are reported in tables. See supplementary materials for a full breakdown of top-line results.

### PPE

Of HCWs who had been to their place of work in the last six weeks (n=831), 80.0% (n=665, 95% CI [77.3 to 82.8]) reported completely adhering to use of PPE the most recent time they were at work.

Complete adherence to use of PPE was associated with older age (table 1). Poorer adherence was associated with having increased face-to-face contact with patients or service users as part of your job. Adherence to use of PPE was poorer in HCWs who reported “rarely” or “often” being in contact with patients with COVID-19, and in those who were never in contact with patients who had COVID-19 but who worked closely with staff who had regular contact with patients who had COVID-19. For these analyses, the reference category was never being in contact with patients with COVID-19 or with staff who had close contact with patients who had COVID-19.

**Table 1.** Participant personal and occupational characteristics, by adherence to use of PPE in the workplace.

| Participant characteristics | Level | Did not completely adhere to use of PPE<br>n=166, n (%) | Completely adherent to use of PPE<br>n=665, n (%) | Odds ratio<br>(95% CI) | Adjusted odds ratio<br>(95% CI)† |
| --- | --- | --- | --- | --- | --- |
| Sex | Male | 48 (20.6) | 185 (79.4) | Reference | Reference |
|  | Female | 118 (19.7) | 480 (80.3) | 1.06 (0.73 to 1.54) | 1.16 (0.77 to 1.74) |
| Age | 18 to 34 years | 31 (35.6) | 56 (64.4) | Reference | Reference |
|  | 35 to 44 years | 39 (22.4) | 135 (77.6) | 1.94 (1.11 to 3.41)* | 1.69 (0.92 to 3.09) |
|  | 45 to 54 years | 46 (18.0) | 209 (82.0) | 2.55 (1.49 to 4.39)** | 2.00 (1.12 to 3.58)* |
|  | 55 years and over | 49 (15.6) | 266 (84.4) | 3.03 (1.78 to 5.16)** | 2.64 (1.49 to 4.66)** |
| Region of place of work ‡ | North East | 9 (23.1) | 30 (76.9) | Reference | Reference |
|  | North West | 22 (21.6) | 80 (78.4) | 1.05 (0.43 to 2.54) | 1.12 (0.44 to 2.86) |
|  | Yorkshire and the Humber | 11 (16.2) | 57 (83.8) | 1.51 (0.56 to 4.08) | 1.77 (0.63 to 5.02) |
|  | East Midlands | 8 (14.0) | 49 (86.0) | 1.8 (0.62 to 5.22) | 1.78 (0.58 to 5.45) |
|  | West Midlands | 13 (21.0) | 49 (79.0) | 1.08 (0.41 to 2.85) | 0.98 (0.35 to 2.74) |
|  | East of England | 17 (32.1) | 36 (67.9) | 0.62 (0.24 to 1.59) | 0.75 (0.28 to 2.04) |
|  | London | 15 (21.7) | 54 (78.3) | 1.01 (0.39 to 2.59) | 0.89 (0.33 to 2.45) |
|  | South East | 27 (23.9) | 86 (76.1) | 0.93 (0.39 to 2.21) | 0.86 (0.34 to 2.16) |
|  | South West | 13 (14.4) | 77 (85.6) | 1.73 (0.66 to 4.49) | 1.71 (0.63 to 4.68) |
|  | Wales | 13 (25.0) | 39 (75.0) | 0.86 (0.32 to 2.28) | 0.76 (0.27 to 2.17) |
|  | Scotland | 14 (13.0) | 94 (87.0) | 1.93 (0.75 to 4.92) | 2.12 (0.8 to 5.67) |
|  | Northern Ireland | 5 (26.3) | 14 (73.7) | 0.86 (0.23 to 3.17) | 0.99 (0.25 to 3.87) |
| Sector ‡ | Private | 43 (22.8) | 146 (77.2) | Reference | Reference |
|  | Public | 123 (19.1) | 520 (80.9) | 1.24 (0.84 to 1.84) | 1.10 (0.71 to 1.69) |
| Work setting ‡ | Pharmacy, dentist, opticians, clinical commissioning group, mental health trust/service, community services, local authority, school, university, other | 36 (15.8) | 192 (84.2) | Reference | Reference |
|  | NHS hospital, private hospital/ clinic, GP surgery/health centre, walk-in centre, ambulance trust/service, care home | 130 (21.6) | 473 (78.4) | 0.69 (0.46 to 1.03) | 0.70 (0.45 to 1.08) |
| Face-to-face contact with patients/service users ‡ | No | 4 (2.8) | 140 (97.2) | Reference | Reference |
|  | Yes, occasionally | 13 (9.9) | 118 (90.1) | 0.25 (0.08 to 0.80)* | 0.25 (0.08 to 0.81)* |
|  | Yes, frequently | 150 (26.9) | 407 (73.1) | 0.07 (0.03 to 0.21)** | 0.07 (0.02 to 0.21)** |
| Occupational group ‡ | Allied health professionals/healthcare scientists/scientific and technical, public health/health improvement, commissioning managers/support staff, wider healthcare team (including admin & clerical, HR, finance, IT, facilities and maintenance), general management, other occupational group | 48 (14.8) | 277 (85.2) | Reference | Reference |
|  | Medical and dental, ambulance, registered nurses and midwives, nursing or healthcare assistants, social care | 75 (23.7) | 242 (76.3) | 0.56 (0.38 to 0.84)* | 0.74 (0.46 to 1.18) |
| Frequency of contact with patients with COVID-19, or staff who worked closely with patients with COVID-19 ‡ | I am never in contact myself with patients who have COVID-19 or anyone who has regular contact with patients who have COVID-19 | 29 (10.3) | 253 (89.7) | Reference | Reference |
|  | I am never in contact myself with patients who have COVID-19 but work closely with staff who have regular contact with patients who have COVID-19 | 21 (15.3) | 116 (84.7) | 0.63 (0.34 to 1.15) | 0.37 (0.19 to 0.74)* |
|  | I am rarely in contact myself with patients who have COVID-19 | 44 (28.0) | 113 (72.0) | 0.30 (0.18 to 0.50)** | 0.36 (0.20 to 0.63)** |
|  | I am sometimes in contact myself with patients who have COVID-19 | 49 (28.8) | 113 (72.0) | 0.28 (0.17 to 0.47)** | 0.43 (0.25 to 0.77)* |
|  | I am often in contact myself with patients who have COVID-19 | 23 (27.1) | 62 (72.9) | 0.31 (0.17 to 0.57)** | 0.55 (0.28 to 1.10) |

\* $p \leq .05$
\*\* $p \leq .002$ (applying Bonferroni correction)
† Adjusting for sex, age, region of place of work, sector, work setting, and face-to-face contact with patients or service users.
‡ The number of valid cases in the table is different from the total count due to the use of weighted data and rounding errors.

There was evidence that workplace environment was associated with adherence to use of PPE. Adherence was associated with having been given all the correct PPE needed to do one’s job; having enough information about what PPE to use and when to use it; and receiving adequate health and safety training at work during the COVID-19 pandemic (table 2).

**Table 2.**
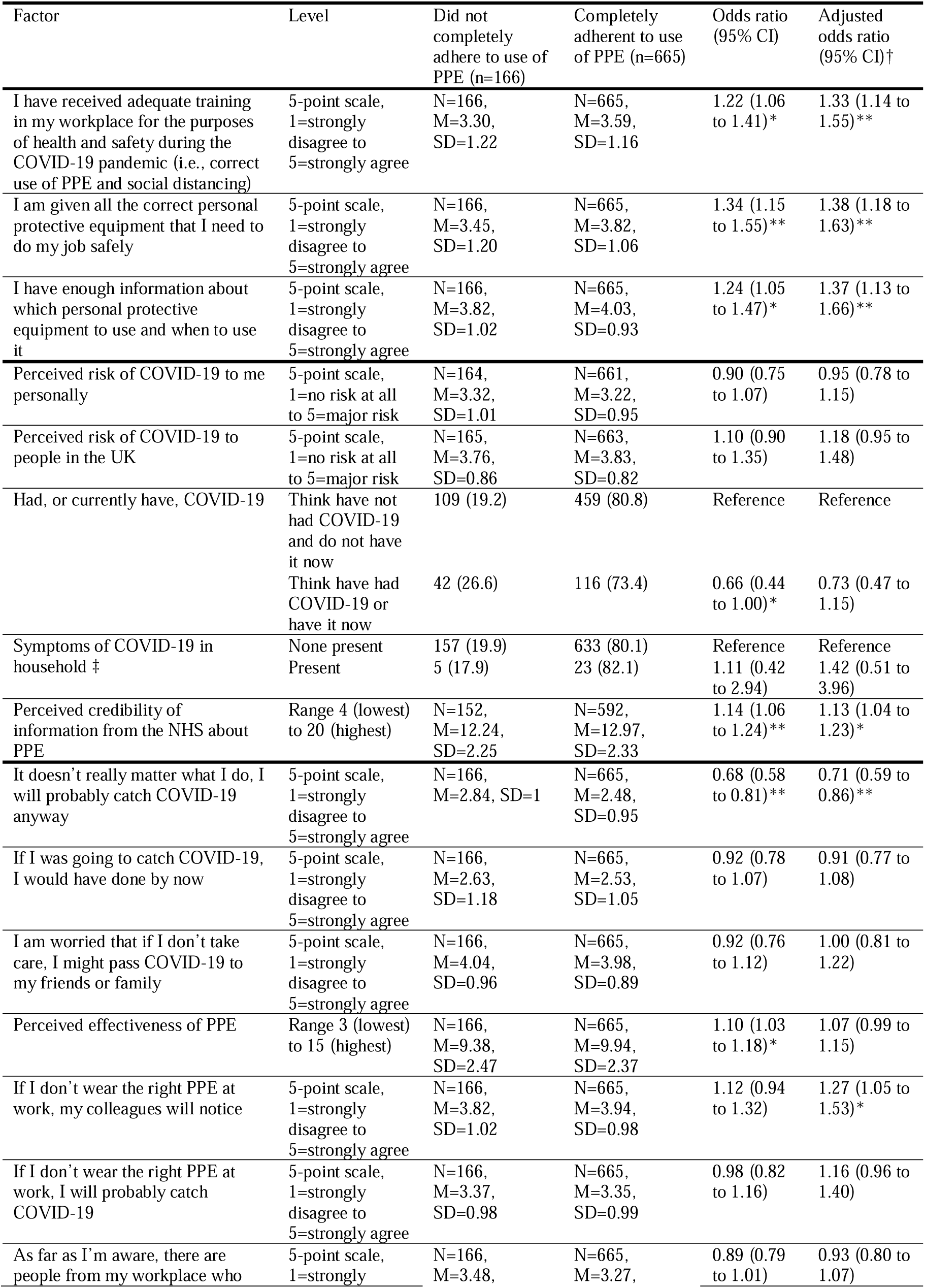
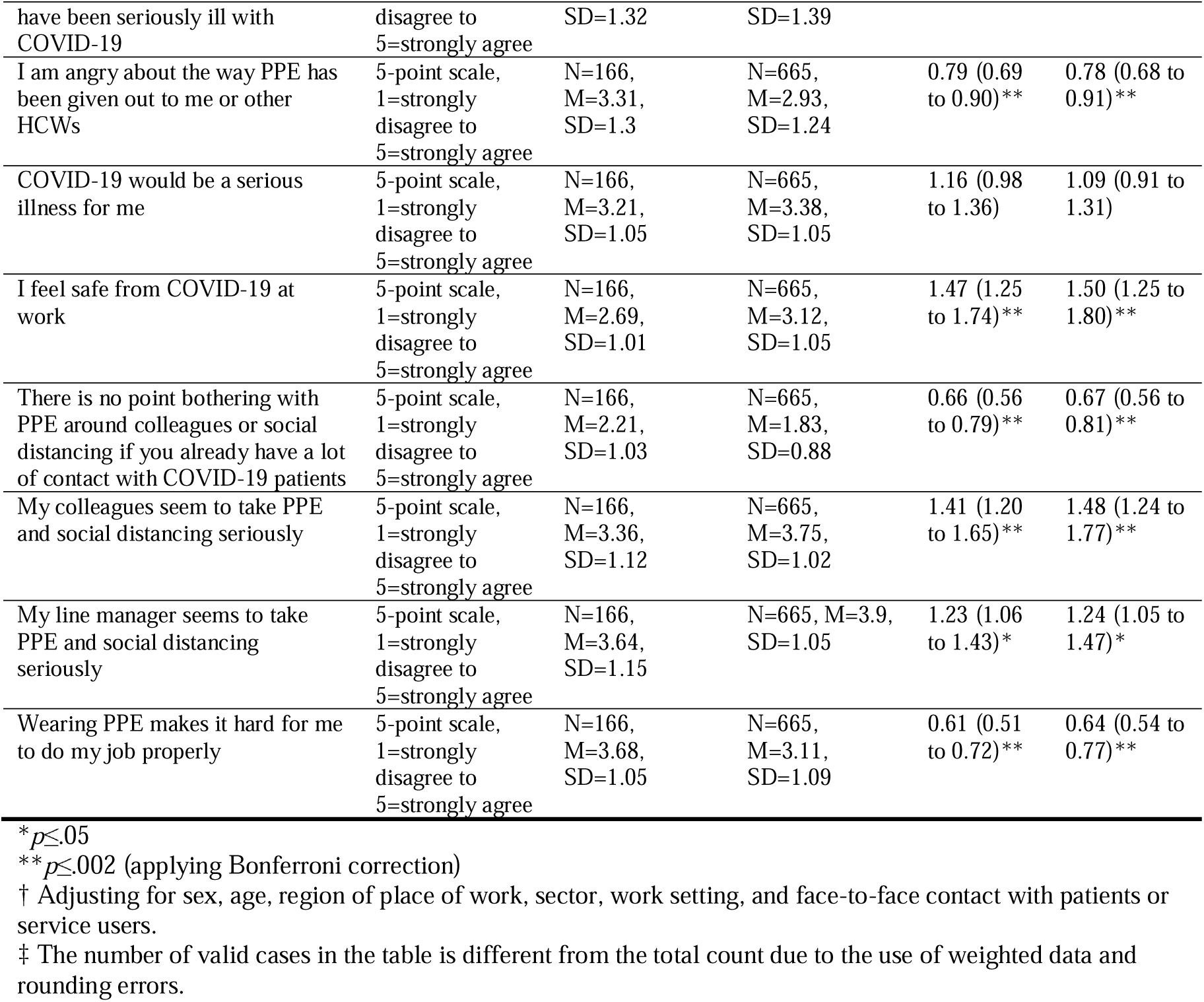
Associations between adherence to use of PPE in the workplace and work environment and psychological and situational factors.

Some psychological factors were associated with complete adherence to use of PPE. Adherence was associated with: thinking that your colleagues and line manager take PPE and social distancing seriously; thinking that your colleagues would notice if you did not wear the right PPE at work; feeling safe from COVID-19 at work; and increased perceived credibility of information from the NHS about PPE (table 2). Lower adherence was associated with agreeing that: wearing PPE makes it difficult to do one’s job; there is no point bothering with PPE or social distancing if you have a lot of contact with patients with COVID-19; thinking that you will probably catch COVID-19 anyway no matter what you do; and being angry about the way PPE had been given out to you or other HCWs.

There was no single source of information about PPE that was preferred by a majority of participants. Instead the most common preferred sources to receive updates about PPE from were: workplace email circulars (32%); line managers (19%); workplace team meetings (15%); infection prevention and control teams (13%; see supplementary materials for full results).

### Hand hygiene

Of HCWs who had been to their place of work in the last six weeks, 67.8% (n=564, 95% CI [64.6 to 71.0]) reported washing their hands every time they needed to during the most recent time they were at work.

Better hand hygiene in the workplace was associated with working in the public sector (aOR 1.50, 95% CI [1.04 to 2.15]) and working in the South East (compared to the North East aOR 2.53, 95% CI [1.13 to 5.66]; full results tables in supplementary materials). Compared to HCWs who never had contact with patients with COVID-19 or with anyone who had regular contact with patients with COVID-19, those who were sometimes in contact with patients with COVID-19 had poorer hand hygiene (aOR 0.63, 95% CI [0.40 to 0.99]).

There was little evidence that workplace related factors were associated with hand hygiene; better hand hygiene was only associated with easy-access handwashing facilities being available when arriving at work (aOR 1.29, 95% CI [1.09 to 1.53]).

There was little evidence that psychological and situational factors were associated with hand hygiene, with better hand hygiene being associated with being worried that you might pass COVID-19 to friends and family if you did not take care (aOR 1.18, 95% CI [1.00 to 1.39]).

### Physical distancing

Of HCWs who had been to their place of work in the last six weeks, 74.7% (n=621, 95% CI [71.7 to 77.7]) reported that they had come into close contact with a colleague the most recent time they were at work.

Close contact with colleagues in the workplace was associated with working in the public sector and in a clinical setting, and having frequent contact with patients with COVID-19 (table 3). Compared to HCWs who never had contact with patients with COVID-19 or with anyone who has regular contact with patients with COVID-19, close contact was associated with: not being in contact with patients with COVID-19 but working closely with staff who have regular contact with patients with COVID-19, and with often being in contact with patients with COVID-19.

**Table 3.** Participant personal and occupational characteristics, by close contact in the workplace.

| Participant characteristics | Level | Were not in close contact with colleagues in the workplace (n=210) | Were in close contact with colleagues in the workplace (n=621) | Odds ratio (95% CI) | Adjusted odds ratio (95% CI)<sup>†</sup> |
| --- | --- | --- | --- | --- | --- |
| Sex | Male | 63 (27.0) | 170 (73.0) | Reference | Reference |
|  | Female | 147 (24.6) | 451 (75.4) | 1.13 (0.80 to 1.59) | 0.94 (0.66 to 1.36) |
| Age | 18 to 34 years | 14 (17.2) | 72 (82.8) | Reference | Reference |
|  | 35 to 44 years | 35 (20.1) | 139 (79.9) | 0.86 (0.44 to 1.67) | 0.97 (0.49 to 1.94) |
|  | 45 to 54 years | 72 (28.2) | 183 (71.8) | 0.55 (0.30 to 1.02) | 0.63 (0.34 to 1.20) |
|  | 55 years and over | 88 (27.9) | 227 (72.1) | 0.55 (0.30 to 1.01) | 0.66 (0.36 to 1.24) |
| Region of place of work <sup>‡</sup> | North East | 8 (20.5) | 31 (79.5) | Reference | Reference |
|  | North West | 28 (27.5) | 74 (72.5) | 0.64 (0.26 to 1.59) | 0.66 (0.26 to 1.67) |
|  | Yorkshire and the Humber | 21 (30.9) | 47 (69.1) | 0.53 (0.21 to 1.37) | 0.47 (0.18 to 1.25) |
|  | East Midlands | 13 (22.8) | 44 (77.2) | 0.85 (0.31 to 2.34) | 0.79 (0.28 to 2.24) |
|  | West Midlands | 16 (25.8) | 46 (74.2) | 0.71 (0.27 to 1.89) | 0.81 (0.29 to 2.23) |
|  | East of England | 13 (24.5) | 40 (75.5) | 0.73 (0.27 to 2) | 0.69 (0.25 to 1.96) |
|  | London | 18 (26.5) | 50 (73.5) | 0.66 (0.26 to 1.72) | 0.71 (0.26 to 1.90) |
|  | South East | 25 (22.1) | 88 (77.9) | 0.84 (0.34 to 2.07) | 0.87 (0.34 to 2.22) |
|  | South West | 21 (23.3) | 69 (76.7) | 0.79 (0.31 to 1.99) | 0.85 (0.32 to 2.21) |
|  | Wales | 10 (19.2) | 42 (80.8) | 0.99 (0.35 to 2.81) | 1.06 (0.36 to 3.11) |
|  | Scotland | 33 (30.6) | 75 (69.4) | 0.55 (0.23 to 1.34) | 0.56 (0.22 to 1.41) |
|  | Northern Ireland | 4 (21.1) | 15 (78.9) | 0.97 (0.24 to 3.91) | 0.90 (0.22 to 3.72) |
| Sector <sup>‡</sup> | Private | 74 (39.2) | 115 (60.8) | Reference | Reference |
|  | Public | 137 (21.3) | 506 (78.7) | 2.39 (1.69 to 3.38)** | 2.43 (1.66 to 3.56)** |
| Place of work <sup>‡</sup> | Pharmacy, dentist, opticians, clinical commissioning group, mental health trust/service, community services, local authority, school, university, other | 81 (35.4) | 148 (64.6) | Reference | Reference |
|  | NHS hospital, private hospital/ clinic, GP surgery/health centre, walk-in centre, ambulance trust/service, care home | 130 (21.6) | 473 (78.4) | 1.99 (1.43 to 2.78)** | 1.65 (1.16 to 2.35)* |
| Face-to-face contact with patients/service users <sup>‡</sup> | No | 40 (28.0) | 103 (72.0) | Reference | Reference |
|  | Yes, occasionally | 45 (34.4) | 86 (65.6) | 0.76 (0.45 to 1.26) | 0.82 (0.48 to 1.39) |
|  | Yes, frequently | 125 (22.5) | 431 (77.5) | 1.35 (0.89 to 2.04) | 1.68 (1.08 to 2.62)* |
| Occupational group <sup>‡</sup> | Allied health professionals/healthcare scientists/scientific and technical, public health/health improvement, commissioning managers/support staff, wider healthcare team (including admin & clerical, HR, finance, IT, facilities and maintenance), general management, other occupational group | 78 (24.0) | 247 (76.0) | Reference | Reference |
|  | Medical and dental, ambulance, registered nurses and midwives, nursing or healthcare assistants, social care | 58 (18.3) | 259 (81.7) | 1.40 (0.96 to 2.05) | 1.50 (0.96 to 2.35) |
| Frequency of contact with patients with COVID-19, or staff who worked closely with patients with COVID-19 ‡ | I am never in contact myself with patients who have COVID-19 or anyone who has regular contact with patients who have COVID-19 | 100 (35.5) | 182 (64.5) | Reference | Reference |
|  | I am never in contact myself with patients who have COVID-19 but work closely with staff who have regular contact with patients who have COVID-19 | 31 (22.6) | 106 (77.4) | 1.90 (1.19 to 3.04)* | 1.68 (1.02 to 2.77)* |
|  | I am rarely in contact myself with patients who have COVID-19 | 35 (22.4) | 121 (77.6) | 1.90 (1.21 to 2.97)* | 1.59 (0.98 to 2.57) |
|  | I am sometimes in contact myself with patients who have COVID-19 | 33 (19.4) | 137 (80.6) | 2.30 (1.46 to 3.61)** | 1.61 (0.96 to 2.67) |
|  | I am often in contact myself with patients who have COVID-19 | 11 (12.8) | 75 (87.2) | 3.74 (1.90 to 7.36)** | 2.43 (1.17 to 5.05)* |

\* $p \leq .05$
\* $p \leq .002$ (applying Bonferroni correction)
† Adjusting for sex, age, region of place of work, sector, work setting, and face-to-face contact with patients or service users.
‡ The number of valid cases in the table is different from the total count due to the use of weighted data and rounding errors.

There was strong evidence that workplace related factors were associated with adherence to physical distancing in the workplace. Less close contact was reported where people reported that: their workplace was designed to make it easy for them to stay 2 metres away from other people; they had received adequate health and safety training at work during the COVID-19 pandemic; and their workplace had clear markings to help you stay 2 metres away from other people (table 4). Perceived ease of physical distancing in the workplace was associated with less close contact with colleagues in the workplace.

**Table 4.**
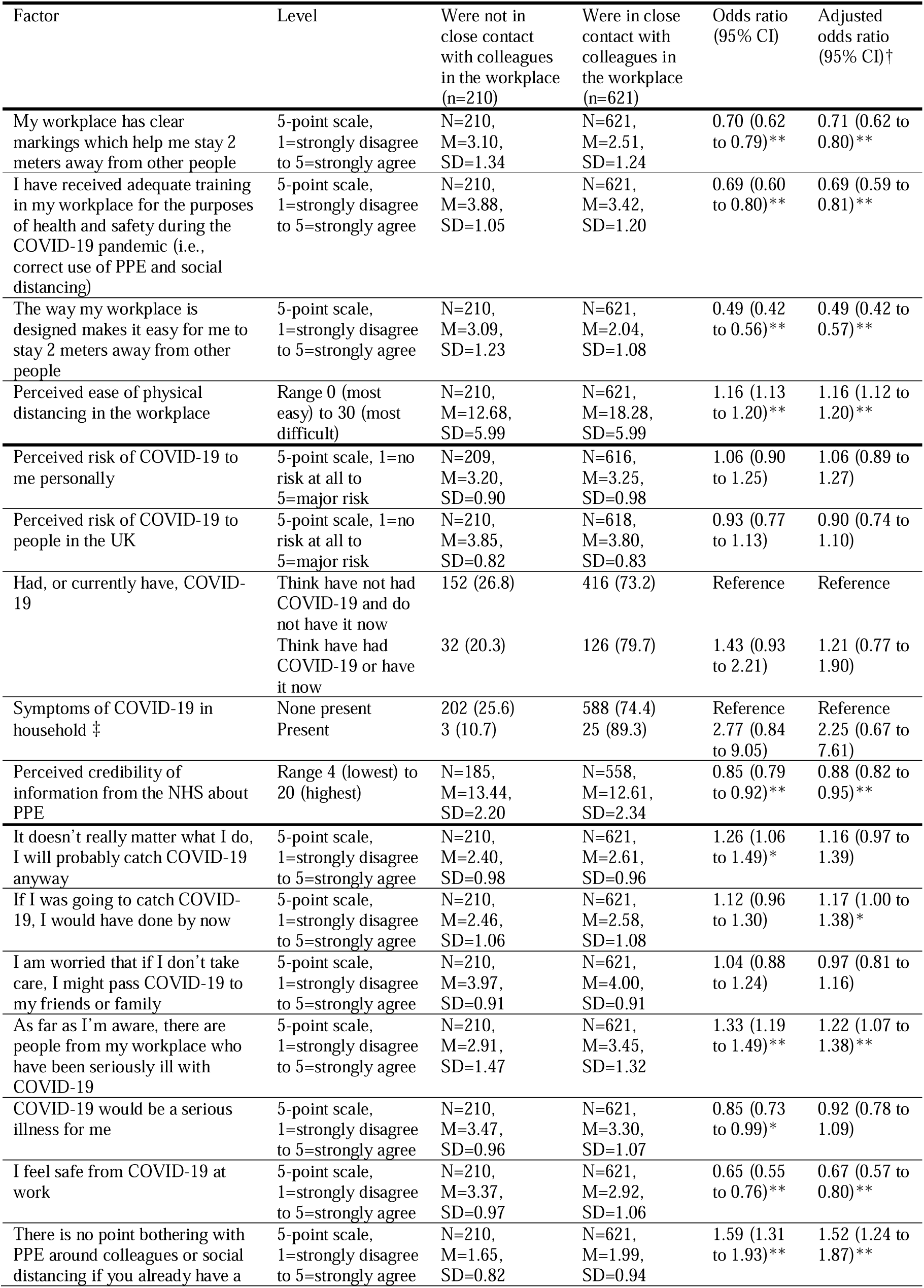
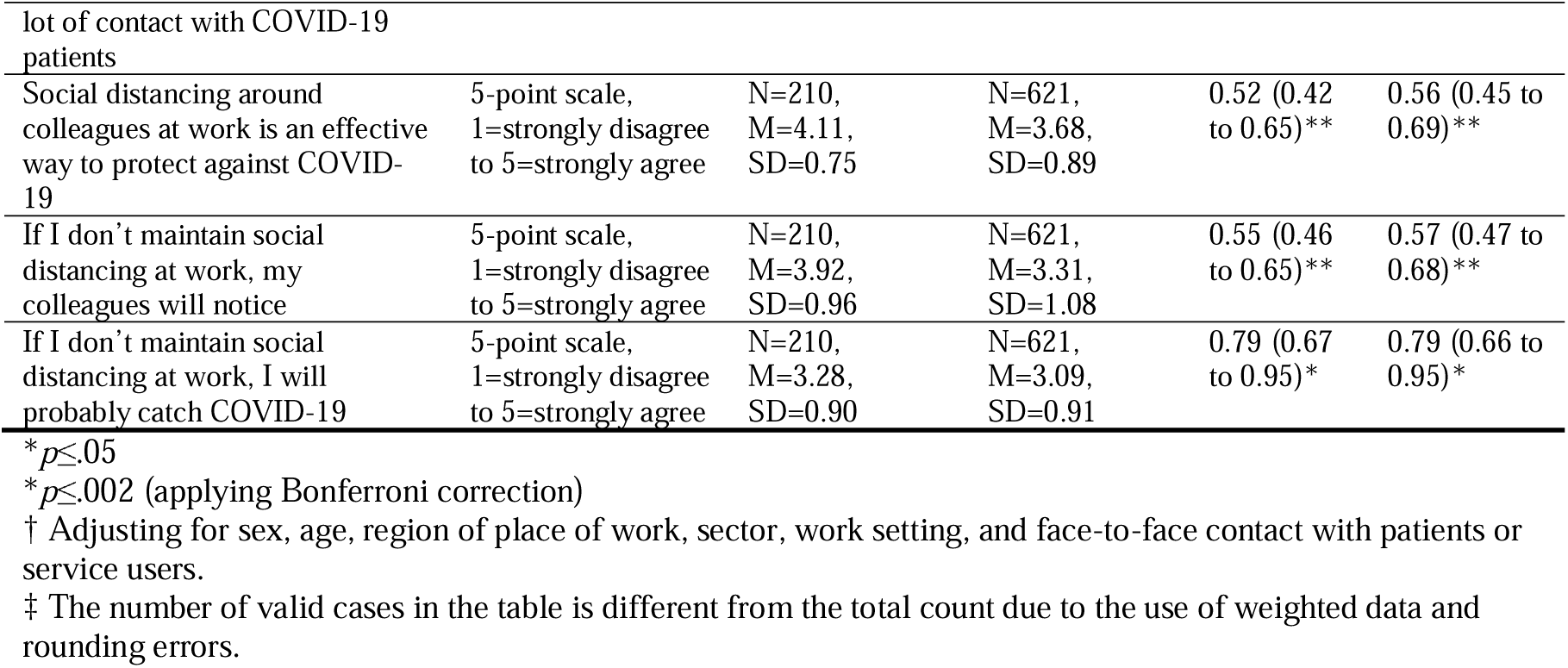
Associations between close contact in the workplace and work environment and psychological and situational factors.

There was good evidence that psychological factors were associated with adherence to physical distancing in the workplace. Greater close contact with colleagues in the workplace was associated with thinking that there is no point bothering with PPE or social distancing if you have a lot of contact with patients with COVID-19; being aware of others in your workplace who had been seriously ill from COVID-19; and thinking that if you were going to catch COVID-19, you would have done so by now (table 4). Less close contact was associated with thinking that social distancing around colleagues at work was an effective way of preventing the spread of COVID-19; that your colleagues would notice if you did not maintain social distancing; feeling safe from COVID-19 at work; thinking that if you did not maintain social distancing at work, you would probably catch COVID-19; and greater perceived credibility of the NHS.

## DISCUSSION

To the best of our knowledge, this is the first study investigating factors associated with adherence to correct use of PPE, hand hygiene and physical distancing measures in healthcare professionals in the UK during the COVID-19 pandemic. Results indicate that adherence to PPE was imperfect, with 20% of HCWs reporting not adhering. Adherence rates should be taken cautiously as they were derived from self-report measures and it was unclear how representative the sample surveyed was to the total population of UK healthcare professionals.

As expected, our sample reported being more likely to adhere to PPE if it was available to them. A recent report by the National Audit Office found that the supply of PPE from central stocks did not meet requirements modelled based on predictions of the reasonable worst-case scenario.^12^ There was also widespread coverage of PPE shortages by the national media during the period covered by our questions.^13, 14^ Anger about how PPE had been distributed was associated with incomplete adherence to usage guidelines. This result suggests that, whilst considerable national PPE procurement efforts were made, a clear communications strategy about the procurement and distribution of PPE may help increase adherence to PPE among HCWs.

The factors most strongly associated with incomplete adherence to use of PPE were reporting more frequent face-to-face contact with patients or service users, and more frequent contact with patients with COVID-19. This may be a function of the greater number of times when PPE was necessary for HCWs with more frequent patient contact. Higher perceived fatalism of catching COVID-19 was also associated with incomplete adherence to use of PPE. Health fatalism is a known barrier for uptake of other health protective behaviours, such as cancer screening.^15^ Such fatalism may be higher in those with increased exposure to the illness at work. Despite previous mixed evidence for the influence of training on adherence to infection control measures,^6^ we found that adherence to PPE use was higher in those who had received health and safety training during the COVID-19 pandemic and who had higher perceived information sufficiency. Thinking that colleagues and line managers took PPE seriously was also associated with increased complete adherence to use of PPE. Results suggest that training on the correct use of PPE during the pandemic could increase adherence. Training and other communications should aim to increase perceived sense of control over the spread of COVID-19 and decrease fatalism by emphasising that complete adherence to use of PPE can help reduce transmission. This may promote a virtuous circle through social norms. Information about PPE use should be delivered through a variety of routes given the lack of any single, preferred route, and be reinforced by line managers and at team meetings.

Around two-thirds (68%) of HCWs reported washing their hands every time they needed to the most recent time they were at work. Given our use of self-report measures, and probable influence of social desirability and recall bias, this is likely an overestimate. This finding should be concerning to healthcare managers. Contrary to previous research,^7, 8^ few factors were associated with good hand hygiene, and there were no strong associations with any particular factor. However, due to space limitations in the survey, we were unable to ask about the perceived effectiveness of handwashing. Further observational research investigating rates of adherence to and factors associated with hand hygiene in HCWs is urgently needed.

Only one-quarter of HCWs reported physically distancing the most recent time they were at work. While we cannot rely on self-reported data to accurately measure rates of adherence, these findings emphasise how difficult physical distancing is between HCWs in the workplace. Clear environmental cues, design of the workplace to enable physical distancing and perceived ease of keeping a 2-metre distance were strongly associated maintaining physical distancing. Patterns of adherence to physical distancing in the workplace were similar to those for adherence to use of PPE, with lower fatalism about catching COVID-19, higher perceived social norms and greater perceived effectiveness of physical distancing being associated with increased adherence. Adherence to physical distancing measures in the UK general population during the pandemic is also associated with greater perceived social norms and effectiveness of physical distancing.^9^ Re-designing areas so that it is easy to maintain physical distancing, and using clear markings to promote physical distancing are likely to promote adherence.

This study has several limitations. First, rates of adherence should be viewed cautiously due to use of self-report data, which may be influenced by recall and social desirability bias. We anticipate that the reported rates are overestimates of adherence. Observational research is needed to investigate rates of adherence to personal protective behaviours in health and social care settings. Second, while the sample was representative of the NHS workforce by occupational group, and weighted data were used to further increase representativeness of the sample, we do not know if survey respondents are truly representative of the wider HCW population.^16, 17^ However, associations within the data may still provide useful insights.^18^ Third, we used cross-sectional data, limiting our ability to infer causation. Fourth, we gathered limited sociodemographic data from participants, due to space constraints in the survey. This prevented us from being able to further investigate the representativeness of survey respondents to the NHS workforce in terms of sociodemographic factors such as education and ethnicity.

This is the first study investigating adoption of protective measures in healthcare professionals in the UK during the COVID-19 pandemic. While reported rates of adherence should be taken with caution, suboptimal levels of adherence reported are likely to be overestimates of real-world adherence. Helpfully, however, our results also suggest several practical solutions to help keep frontline workers safe (table 5). First, it should be ensured that health and social care professionals have adequate PPE resources. Second, HCWs should be sufficiently trained in protective measures in place during the COVID-19 pandemic. Such training should emphasise personal control over the spread of the virus through correct use of PPE, hand hygiene and physical distancing in the workplace and aim to decrease fatalism of catching COVID-19. Adoption of these protective measures may also increase sense of safety in the workplace. Different to adherence to other protective measures, perceived risk of COVID-19 was not associated with adhering to personal protective behaviours in the workplace. These findings suggest that training should focus on more practical aspects of protective measures rather than the risk of COVID-19. Third, workplaces should be set up to encourage physical distancing by using clear markings. Fourth, a culture of adopting PPE, good hand hygiene and maintaining physical distancing will promote adherence. This may encourage a virtuous cycle and help promote good practice beyond the pandemic period.

**Table 5.** Summary of research findings and their implications for healthcare managers

| Research finding | Implication |
| --- | --- |
| Higher adherence to PPE use and physical distancing in those who have received adequate workplace health and safety training during the COVID-19 pandemic | HCWs should receive training on protective measures in place during the COVID-19 pandemic |
| Lower adherence to PPE use and physical distancing in those with higher fatalism about COVID-19 | Training should aim to decrease fatalism about COVID-19, by emphasising personal control over the spread of the virus through correct use of PPE, hand hygiene and physical distancing in the workplace |
| Higher adherence to PPE use and physical distancing in those with higher perceived social norms | A culture of adopting personal protective behaviours will encourage better adherence |
| Lower adherence to PPE in those who did not have adequate PPE resources | Adequate PPE should be given to all HCWs |
| Higher adherence to physical distancing in those whose workplaces have clear markings and make it easy to maintain physical distance | Workplaces should be set up to encourage physical distancing by using clear markings |
| No clear associations between contact with COVID-19 patients and increased adherence to protective measures in the workplace | Training should be delivered to all staff, tailored to the use of protective measures appropriate to their occupational goal |

Fifth, there were no clear associations between contact with COVID-19 patients and increased adherence to protective measures in the workplace. Therefore, training should be delivered to all staff, tailored to the use of protective measures appropriate to their occupational role.

## Data Availability

Anonymised data will be made available upon reasonable request.

## AUTHOR CONTRIBUTION STATEMENT

The study was conceptualised by NG and GJR. LS completed all analyses, using data from YouGov Plc. All authors contributed to, and approved, the final manuscript. For any enquiries about the data in this report please contact King’s College London.

